# Normal growth curve of choroid plexus in children: Implications for assessing hydrocephalus due to choroid plexus hyperplasia

**DOI:** 10.1101/2023.05.10.23289689

**Authors:** Hiroaki Hashimoto, Osamu Takemoto, Keisuke Nishimoto, Gento Moriguchi, Motoki Nakamura, Yasuyoshi Chiba

## Abstract

**Objective:** Pediatric hydrocephalus requires evaluation while accounting for growth of intracranial structures, but information on choroid plexus growth in children is lacking. This study aimed to create normal growth curves for intracranial volume, choroid plexus volume, and lateral ventricles volume and assess objectively the degree of hydrocephalus due to choroid plexus hyperplasia (CPH) and the effect of surgeries.

**Methods:** This retrospective study analyzed head computed tomography (CT) scans of pediatric patients with head trauma from Osaka Women’s and Children’s Hospital between April 2006 and April 2023. The study segmented the intracranial volume, choroid plexus, and lateral ventricles and calculated their volumes. The study also calculated correlation coefficients among the three parameters. Patients aged 0 to 10 years were divided into 15 age-related clusters and mean and standard deviation (SD) values were measured in each cluster. Growth curves were created by plotting mean values sequentially. Volume obtained from patients with CPH were z-normalized using mean and SD values and compared.

**Results:** A total of 222 CT scans (91 from females) were analyzed, and positive correlations were observed among intracranial volume, choroid plexus volume, and lateral ventricles volume, with the strongest correlation between choroid plexus and lateral ventricles volumes. The growth rate of intracranial volume was rapid until approximately 20 months of ages, while those of choroid plexus and lateral ventricles volume increased rapidly by approximately one year of age.

After that, the volume reached plateau at 1.5 mL and 10mL in choroid plexus and lateral ventricles volume, respectively. Three patients with CPH were enrolled and quantitatively evaluated by the z-normalized volume (z.v.). Notable abnormal volume of choroid plexus (ranged z.v. 26.00 – 45.85) and lateral ventricles (ranged z.v.40.93 – 123.56) were observed. In two patients, z.v. lateral ventricles volumes improved after surgical interventions. Choroid plexus volume reduced by approximately 20% (from z.v. 45.85 to z.v. 36.95) after bilateral endoscopic plexus coagulation in one patient.

**Conclusions:** This study provides normal growth curves for intracranial volume, choroid plexus volume, and lateral ventricles volume. Knowledge of normal values enables objective assessment of abnormal values related to hydrocephalus and choroid plexus disease such as CPH.

## Introduction

Hydrocephalus is a common disorder in both pediatric and adult neurosurgical fields. Although reliable incidence figures for adult hydrocephalus are absent in the literature, the incidence of congenital hydrocephalus in the United States/Canada has been estimated at 68 per 100,000 births.^1^ Hydrocephalus has been classically defined as any increase in cerebrospinal fluid (CSF) within the skull ^2^, or as an active distension of the ventricular system of the brain resulting from inadequate passage of CSF from its point of production within the cerebral ventricles to its point of absorption into the systemic circulation.^3^ Therefore, there appears to be a close relationship between ventricular size and CSF production, and the choroid plexus was experimentally demonstrated as the origin of CSF production in the beginning of the 1900s.^4^ In the neuroscience field, the production and directional flow of CSF have been highlighted along with the glymphatic concept.^5^ In clinical situations, rare cases of choroid plexus papilloma or choroid plexus hyperplasia (CPH) show hydrocephalus due to an overproduction of CSF, which are treated by shunt procedures or microsurgical removal.^6^

Normal growth curves related to head circumference are well-established tools for physicians to assess abnormal growth patterns. Normal growth curves of intracranial volume have been reported by volumetry using computed tomography (CT) imaging,^7^ and those of cerebral ventricular volume have been revealed by volumetry using magnetic resonance imaging (MRI).^8^ Normal thickness of the choroid plexus has been reported in child population^9^ and adult population^10^, and aging in adult population was associated with increases in choroid plexus volume.^11^ Recently, the volume of the choroid plexus has been highlighted in disease such as psychosis spectrum,^12^ Alzheimer disease,^13^ multiple sclerosis,^14^ and stroke.^15^ However, the normal figure of child choroid plexus volume and how it grows in the pediatric population remains unclear. Additionally, knowledge about the relationship among pediatric intracranial volume, choroid plexus volume, and intracranial ventricular volume has been lacking.

In this study, we enrolled pediatric patients with head trauma who were healthy before suffering from head injury and who underwent head CT to measure volumes of intracranial volume, choroid plexus volume, and lateral ventricles volume. We assessed the relationship among them and created growth curves. Quantitative assessment of these parameters enables us to evaluate hydrocephalus more objectively. Finally, we measured volumes of these parameters obtained from patients with CPH, and assessed the degree of abnormality and the effect of surgical procedures.

## Methods

### Patients and study setting

In this retrospective study, we enrolled patients who underwent CT imaging due to head trauma at Osaka Women’s and Children’s Hospital between April 2006 and April 2023. As the significant dose-response relationship was observed between pediatric CT-related radiation exposure and brain cancer,^16^ our institution follows guidelines to determine whether CT is performed on children with head trauma.^17,18^ Patients were excluded based on the following criteria: (1) age ≥11 years, (2) suspicion of abuse, (3) need for craniotomy for decompression within a few days after head injury, (4) cranial depressed fracture requiring recovery operation, (5) presence of complications such as craniosynostosis, tumor, epilepsy, autism, intracranial arachnoid cyst, chromosomal abnormalities, cardiovascular disease, and endocrine disorders etc., and (6) presence of cavum Vergae or cavum septum. Therefore, all subjects were healthy prior to the head injury, and no operations were needed. In some cases, follow-up CT scans were performed due to medical necessity. The Ethics Committee of Osaka Women’s and Children’s Hospital (Izumi, Japan, approval no. 1634) provided ethical approval for this study, which was conducted in accordance with the Declaration of Helsinki guidelines for experiments involving humans. Informed consent was obtained using the opt-out method from our center’s website because of the retrospective and noninvasive nature of the study.

### Data collection

We retrospectively collected CT scans and evaluated medical variables related to patients, including sex and age. For quantitative assessment of intracranial volume, volume of choroid plexus, and volume of lateral ventricles, Digital Imaging and Communications in Medicine (DICOM) CT date were imported to MATLAB R2020b (MathWorks, Natick, MA, USA), and the target areas were segmented manually using the image segmenter app in MATLAB (https://www.mathworks.com/help/images/ref/imagesegmenter-app.html). The width of DICOM slice was 5 mm. Choroid plexus observed in the bilateral ventricles were assessed as a representation of intracranial choroid plexus. The volume of lateral ventricles was measured as the volume of bilateral ventricles. These procedures enabled us to calculate the volume (in milliliter, mL) related to the above parameters, and this method has been used in our other studies.^19,20^

### Statistical analyses

Categorical data were presented as frequencies (percentages). Continuous variables with normal distribution were presented as mean ± standard deviation (SD) while those with non-normal distribution were presented as median with 25%-75% quartiles. Spearman correlation coefficients were calculated to evaluate correlations among parameters.

To account for the uneven distribution of age, we assessed the distribution and defined age-related clusters for adjustment. Mean and 95% confidence intervals (CIs) of parameters were calculated, and differences between age-related clusters or genders were compared by assessing overlapping or non-overlapping CIs. Non-overlapping CIs were considered as an indicator of statistical significance, while overlapping CIs were interpreted as indicating a lack of statistical significance.^21^ Additionally, unpaired T-tests were used for comparison among age-related clusters, and Bonferroni correction was applied for the correction of multiple comparison. The statistical difference between clusters was considered significant at corrected p-values of <0.05.

The measurements of each parameter were plotted according to months after birth, and best-fit logarithmic curves were plotted. Statistical analyses were performed using the Statistical and Machine Learning Toolbox of MATLAB R2020b.

### Choroid plexus hyperplasia

We collected data from patients with CPH who underwent surgeries at our institution between April 2006 and April 2023. Volumes of parameters were calculated and z-normalized by subtracting the mean values and dividing by SD of the corresponding age-related clusters.

### Data availability

All data in this study are available from the corresponding authors upon reasonable request and after additional ethics approval.

## Results

### Baseline characteristics

Data from a total of 222 CT scans performed in 177 patients with head trauma were collected. Of the 222 CT scans, 91 (41.0%) were obtained from females, and the median value of months after birth was 13 (6–42) months. CT scans were performed twice in 41 patients (17 females, 41.5%), and three times in two patients (0 female, 0.0%).

### Correlation analysis

Positive correlations were observed among three parameters, including intracranial volume, volume of choroid plexus (choroid plexus volume), and volume of lateral ventricles (lateral ventricles volume), in all three combinations (Fig.1). Notably, the correlation coefficient between choroid plexus and lateral ventricles volumes reached the highest value, which was significant (r = 0.53, *p* = 2.58 × 10^−17^) (Fig.1C).

**Fig. 1.**
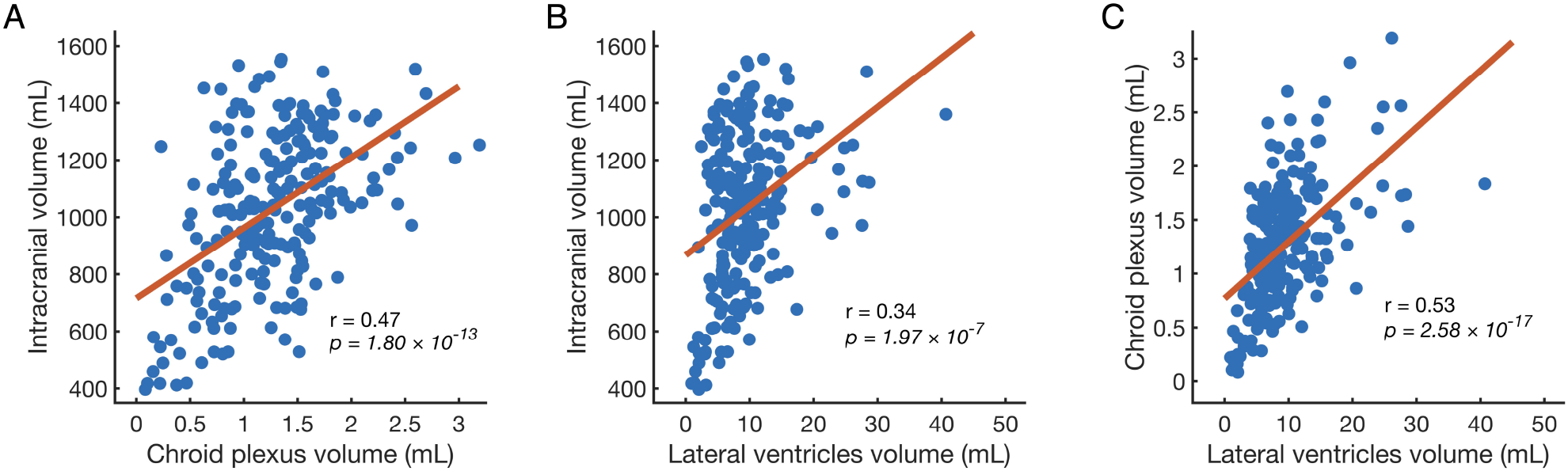
Correlation relations between parameters. Scatter plots between intracranial volume and choroid plexus volume (A), intracranial volume and lateral ventricles volume (B), and choroid plexus and lateral ventricles volume (C) are presented. The regression lines are indicated as red lines and *r* represents the correlation coefficients.

### Age-related clusters

To account for the uneven distribution of the population across different age groups, we categorized the CT scans into fifteen age-related clusters: 0 month, 1 month, 2–3 months, 4–5 months, 6–7 months, 8–9 months, 10–11 months, 1 year-1 (age range 12 to 17 months), 1 year-2 (age range 18 to 23 months), 2 years, 3 years, 4 year, 5–6 years, 7–8 years, and 9–10 years. The number of CT scans performed in each cluster for both males and females is presented in Figure 2. The population distribution between males and females was uneven due to higher frequency of CT scans for head trauma in males compared to female.

**Fig. 2.**
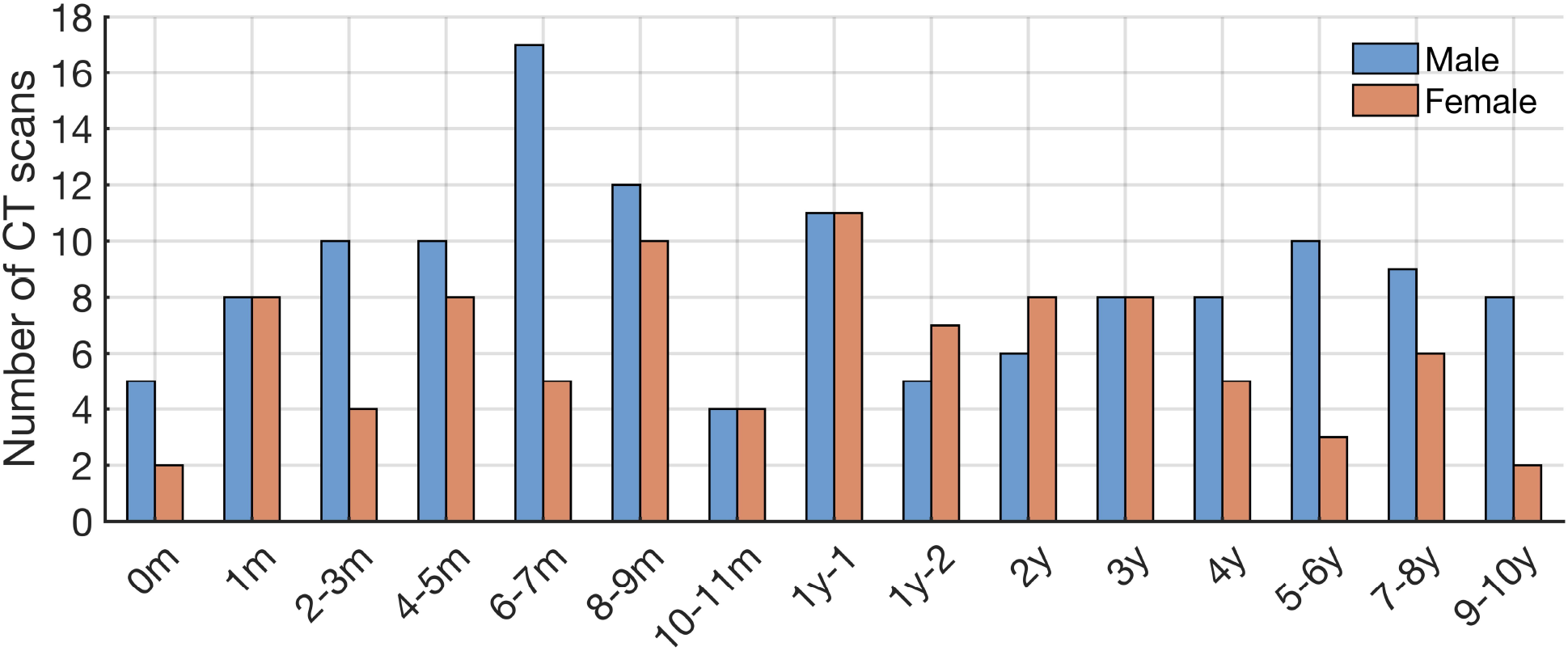
Distribution of population among age-related clusters. Fifteen age-related clusters were defined, ranging from 0 month (m) to 9-10 years(y). The cluster labeled as “1y-1” corresponds to an age range of 12 to 17 months, and the cluster labeled as “1y-2” corresponds to an age range of 18 to 23 months.

### Sequential plots of parameters

The mean values of each cluster, calculated from the total number of CT scans, were plotted sequentially for intracranial volume (Fig.3A), choroid plexus volume (Fig.3B), and lateral ventricles volume (Fig.3C). Intracranial volume consistently increased step by step with each age-related cluster, while both choroid plexus and lateral ventricles volume increased until approximately 1 year of age, after which the subsequent increased slowed down. In intracranial volume, the error bars indicating 95% CIs showed almost non-overlapping among each cluster. However, in choroid plexus and lateral ventricles, the error bars after 1 year of age showed almost overlapping.

In the lower row in Figure 3, plots generated from males and females were shown. Except for 6–7 m and 8–9 m in the intracranial volume (Fig.3D), the error bars of males and females showed overlapping in each cluster. We wanted to note that due to the small sample size of females, 95% CIs indicated negative values (Fig.3E, and 3F). Therefore, we judged that in this study, we could not present any differences among males and females due to small sample size, and in the following analyses, we presented results obtained from the total CT scans.

**Fig. 3.**
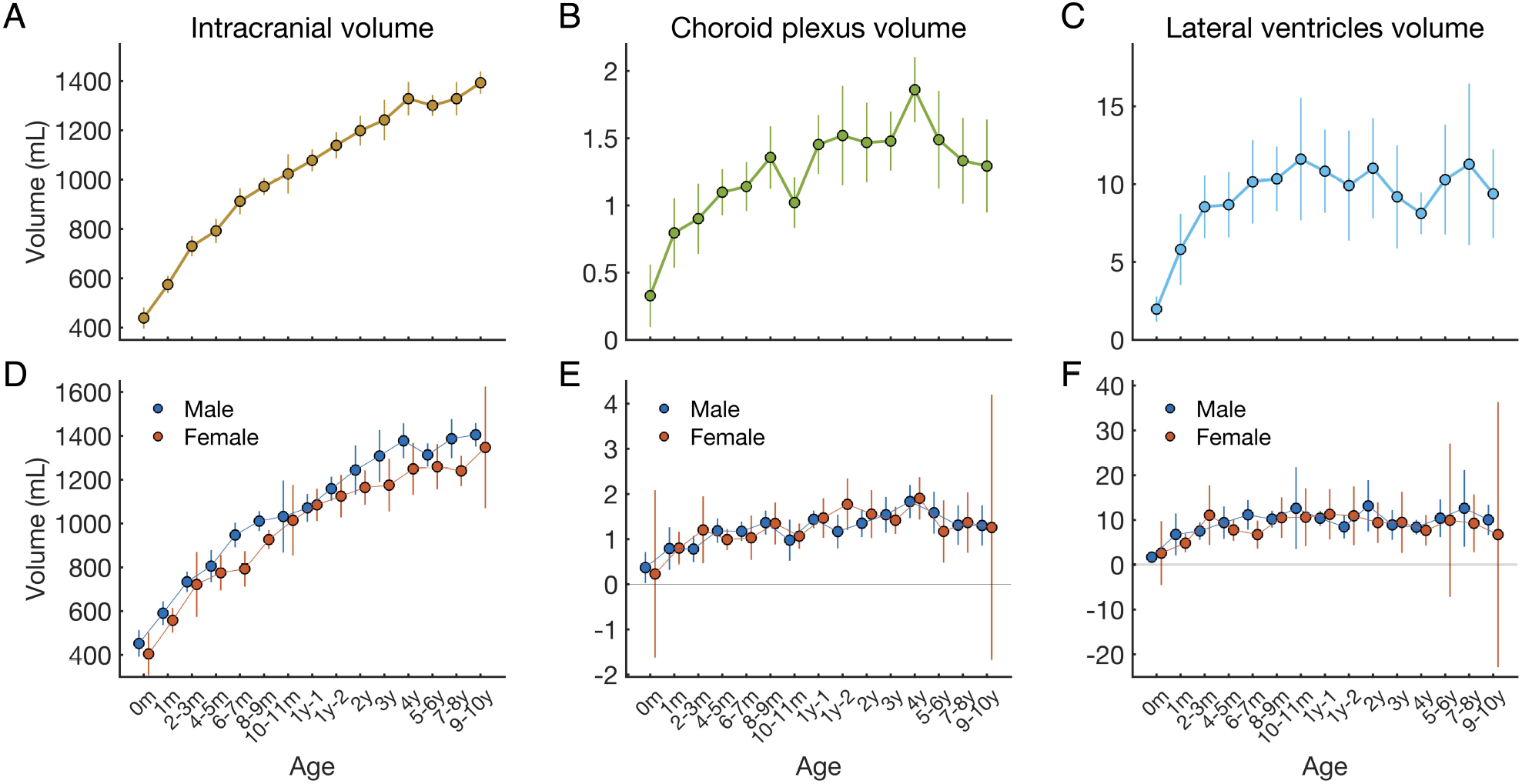
Sequential plots of mean values. The mean values are calculated in each age-related cluster and sequentially plotted among three parameters. The panels of the upper row are obtained from total CT scans (A, B, and C), and the panels of the lower row are obtained from CT scans in males and females (D, E, and F). Error bars indicate 95% confidence intervals.

### Statistical differences among each age-related cluster

To compare the volumes between two different age-related clusters for each parameter, we performed unpaired T-tests. To account for multiple comparisons, we applied Bonferroni correction by multiplying the obtained p-values by 105, which was the total number of combinations (_15_C_2_). The significant differences in volume, with corrected p-values less than 0.05, were presented as a color-scaled matrix (Fig.4). We observed significant differences in intracranial volume between two different age-related clusters below two years of age (Fig.4A). However, except for the combination between 0 months and other age-related clusters, we found almost no significant differences between clusters in choroid plexus and lateral ventricles volume (Fig.4B and 4C).

**Fig. 4.**
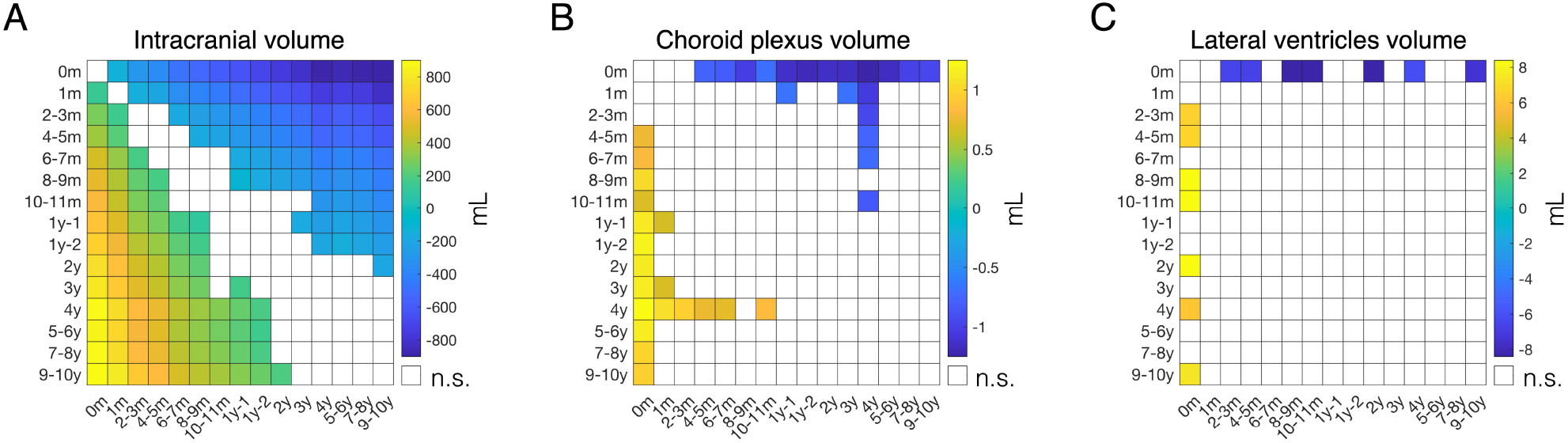
Color-scaled matrix of significant differences in volume. Differences among age-related cluster in intracranial volume (A), choroid plexus volume (B), and lateral ventricles volume (C) are presented as a color-scaled matrix. Positive differences, indicating larger volumes in the longitudinal axis compared to the horizontal axis, are colored in green to yellow. Negative differences, indicating smaller volumes in the longitudinal axis compared to the horizontal axis, are colored in blue. Only differences that showed significancy with corrected p-values <0.05 are colored. n.s., not significant.

### Growth curve

Scatter plots against age in months were used to generate best-fit logarithmic curves; y = 199.17log(x+1) + 491.55 for intracranial volume (Fig.5A), y = 0.19log(x+1) + 0.75 for choroid plexus volume (Fig.5B), and y = 0.91log(x+1) + 6.94 for lateral ventricles volume (Fig.5C). In intracranial volume, the growth rate was faster until approximately 20 months of age, and slowed down after 20 months. However, in choroid plexus and lateral ventricles volume, the growth rate slowed down quickly by approximately 10 months of age.

**Fig. 5.**
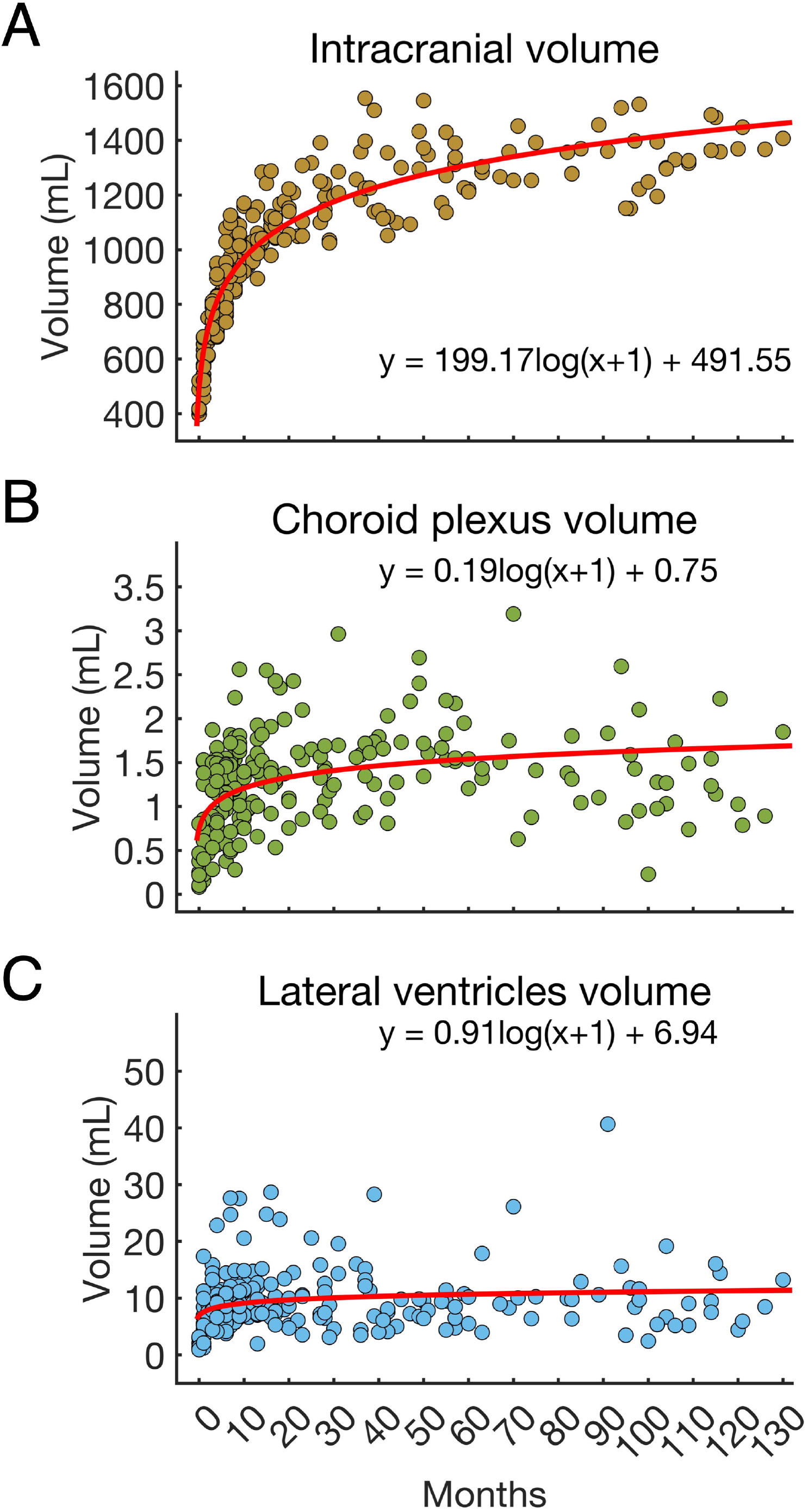
Scatter plots and growth curves. Scatter plots against age in months are presented for intracranial volume (A), choroid plexus volume (B), and lateral ventricles volume (C). The best-fit logarithmic curves are indicated as red curves.

### Cases with CPH

Three cases with CPH were enrolled, and their profiles are presented in Table 1. Two of them required more than two operations due to complications and underwent external ventricular drainage (EVD). As a result, we were able to measure the daily production of cerebrospinal fluid, which were notably over 2,000mL. Additionally, two of the cases had genetic abnormalities, such as trisomy 9p.

**Table 1.**
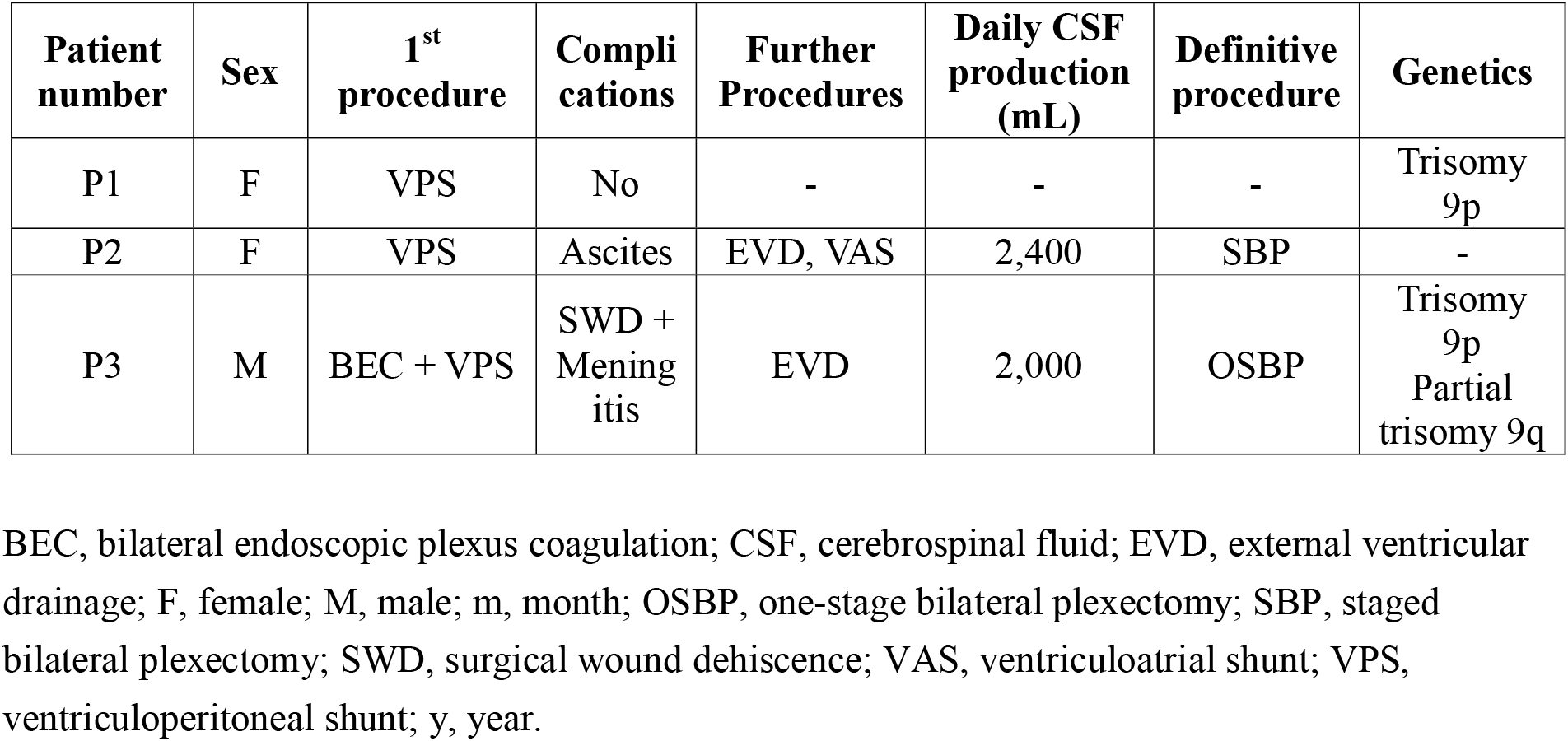
Clinical profile of patients with choroid plexus hyperplasia.

| **Patient number** | **Sex** | **1<sup>st</sup> procedure** | **Complications** | **Further Procedures** | **Daily CSF production (mL)** | **Definitive procedure** | **Genetics** |
| --- | --- | --- | --- | --- | --- | --- | --- |
| P1 | F | VPS | No | - | - | - | Trisomy 9p |
| P2 | F | VPS | Ascites | EVD, VAS | 2,400 | SBP | - |
| P3 | M | BEC + VPS | SWD + Meningitis | EVD | 2,000 | OSBP | Trisomy 9p<br>Partial trisomy 9q |

BEC, bilateral endoscopic plexus coagulation; CSF, cerebrospinal fluid; EVD, external ventricular drainage; F, female; M, male; m, month; OSBP, one-stage bilateral plexectomy; SBP, staged bilateral plexectomy; SWD, surgical wound dehiscence; VAS, ventriculoatrial shunt; VPS, ventriculoperitoneal shunt; y, year.

The volumes of parameters were calculated and z-normalized using mean and SD values obtained from normal age-related clusters (Supplemental Table 1). The results were plotted over the plots calculated from the normal groups (Fig.6A, 6B, and 6C). The z-normalized values of choroid plexus and lateral ventricles volume were notably larger than those of intracranial volume.

**Fig. 6.**
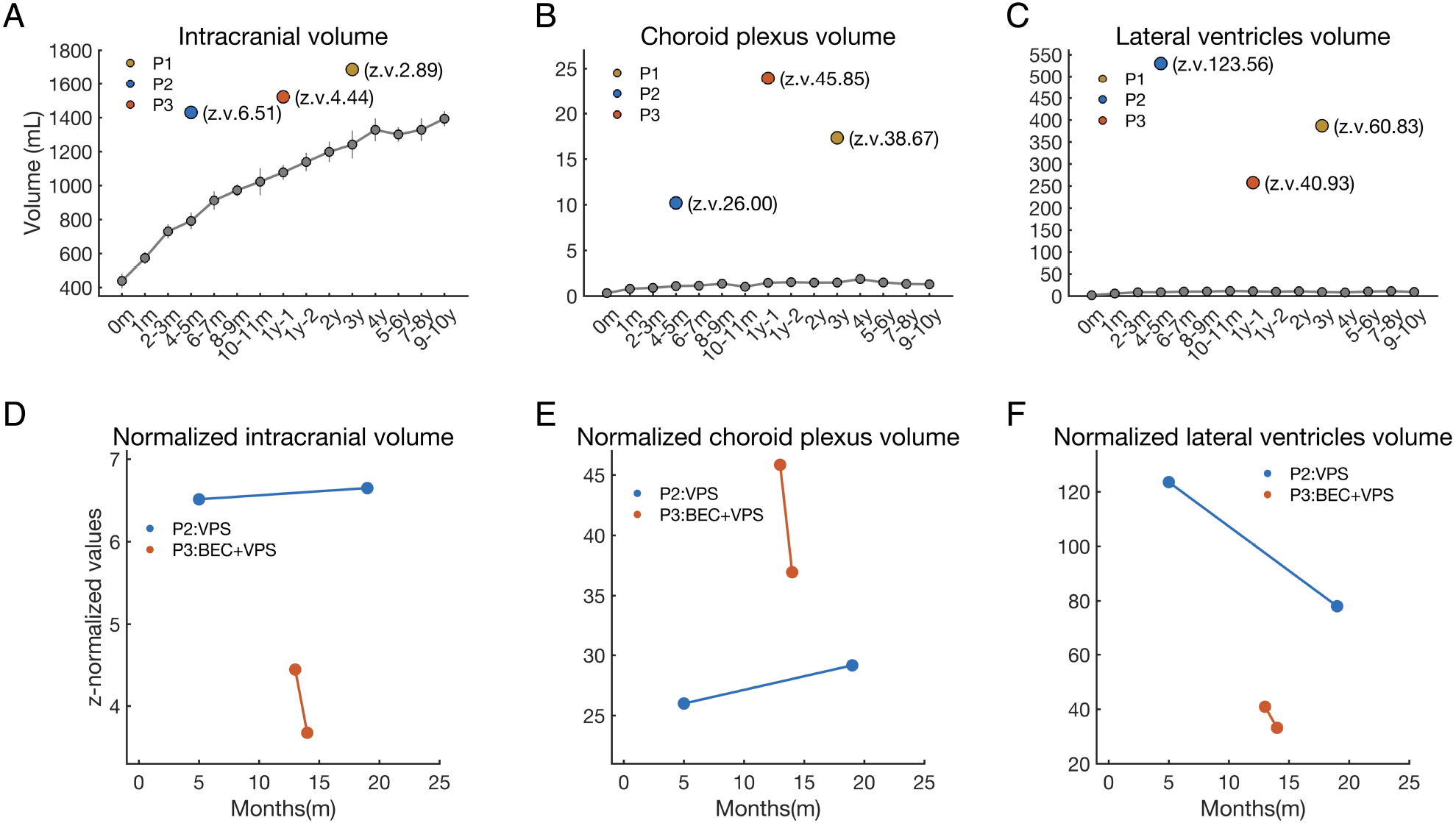
Results from cases with CPH. Volumes calculated from patients with CPH in each parameter are presented in the upper row (A, B, and C) with plots and lines calculated from normal age-related clusters indicated in gray color, similar to the upper panels of Figure 3. The changes of z-normalized values of P2 and P3 in each parameter are presented in the lower row (D, E, and F). BEC, bilateral endoscopic plexus coagulation; VPS, ventriculoperitoneal shunt; z.v., z-normalized values.

Specifically, for P2 and P3, the changes in z-normalized values from the time of initial diagnosis of CPH to the time of EVD were presented in Fig.6D, 6E and 6F. P2 underwent ventriculoperitoneal shunt (VPS) alone, while P3 underwent VPS combined with bilateral endoscopic plexus coagulation (BEC) during these intervals. Therefore, we were able to evaluate the effect of these operations. While the z-normalized intracranial volume of P2 increased, those of P3 decreased. Due to BEC, the z-normalized choroid plexus volume of P3 decreased from 45.85 to 36.95 (19.4% down). In both of P2 and P3, the enlarged lateral ventricles volume improved but remained abnormally large.

## Discussion

Using DICOM images of CT scans obtained from Japanese pediatric patients with head trauma, we generated normal growth curves for intracranial volume, lateral ventricles volume, and choroid plexus volume. We found positive correlations among these parameters, with the strongest correlation observed between choroid plexus and lateral ventricles volume. The growth rate of intracranial volume was rapid until approximately 20 months of age, while the growth rates of choroid plexus and lateral ventricles volume slowed down by approximately one year of age. After that, the volume of choroid plexus and lateral ventricles remained approximately 1.5mL and 10mL, respectively. We utilized z-normalization with mean and SD values obtained from the normal growth curves to evaluate the degree of hydrocephalus caused by CPH. We also assessed the effect of surgical procedures on hydrocephalus in two patients (P2 and P3). We observed a decrease in lateral ventricles volume, indicating an improvement in hydrocephalus, in both patients after surgery. In P3, we also observed a reduction in choroid plexus volume by approximately 20% after BEC.

We stratified the pediatric patients into age-related clusters and calculated their mean, 95% CI, and SD, enabling us to calculate z-normalized values and compare them across different ages. In contrast to adult studies, where choroid plexus volume was corrected by intracranial volume due to head size variability,^11^ the wider head size variability among pediatric patients poses a challenge for a quantitative assessment of pediatric hydrocephalus across different ages. Our study provides normal values, which enable the objective assessment of abnormal values. The actual intracranial volume of P1 was the highest among the three patients, but when z-normalized, it had the lowest value. The normal values of intracranial volume enable quantitative evaluation of abnormal head size resulting from conditions such as hydrocephalus or craniosynostosis, among others. While the Evans’ index, a two-dimensional measurement, has been used to assess normal pressure hydrocephalus, its insufficiency has been noted,^22,23^ and the importance of three-dimensional measurement of the ventricular volume for ventriculomegaly assessment has been emphasized.^8^ Our results on choroid plexus volume can be used to evaluate the abnormal volume of rare cases, such as CPH, choroid plexus papilloma, and choroid plexus carcinoma, which can cause hydrocephalus.^24^

Previous studies have reported growth curves of intracranial volume using thin-slice head CT images to evaluate 106 Japanese pediatric subjects.^7^ Although our study used CT images in 5mm slice width, our growth curves of intracranial volume were similar to those presented in the previous study. In addition, growth curves related to normal cerebral ventricular volume have been also reported, where brain MRI images were analyzed, and 687 MRI images from the University of Michigan database were investigated.^8^ The previous study showed that rapid growth occurred during the first year of life, and the 50th percentile of ventricular volume after one year of age was approximately 10mL, which is consistent with our results. These concordances between previous findings and our results validate our methodology and results. Additionally, it is likely that differences in the growth of these parameters between different races are small.

Although our results did not reveal obvious differences between males and females, it is possible that our sample size, particularly for females, was insufficient to determine whether there is a gender-related difference. The percentage of females in our study was approximately 40%, which may have been lower than that of males given that we enrolled children suffering from head trauma. A previous study investigating head injury in individuals aged 15 to 79 years showed that the population of females was only 23%.^25^ These findings suggest that males have a higher incidence of head trauma compared to females in both pediatric and adult populations.

However, a previous study evaluating adult ventricular volume showed that the effect of sex was statistically non-significant,^11^ and another study evaluating pediatric ventricular volume found that female ventricular volumes were slightly lower than male volumes.^8^ Therefore, we infer that there might be no statistical difference among genders in terms of the growth of intracranial volume, lateral ventricles volume, and choroid plexus volume.

The choroid plexus is a major source of CSF and also secretes a large variety of trophic factors in the CSF that have an impact on the entire brain.^26^ Recently, choroid plexus volume has been associated with neurological disease, with significant increases reported in patients with stroke,^15^ multiple sclerosis,^27^ and psychosis.^12^ In the normal adult population, choroid plexus volume has been reported to increase with age.^11^ The calculated choroid plexus volume from adult control groups was approximately 1.2mL,^15^ and our study also revealed that after one year of age, choroid plexus volume remained approximately 1.5mL. These findings suggest that the growth rate of choroid plexus volume is rapid until approximately one year of age, and that the initial growth of the choroid plexus may be completed by one year of age. After the age of 50 years,^11^ the volume of the choroid plexus may begin to grow again.

The rapid growth observed in choroid plexus volume by one year of age was similar to that observed in lateral ventricles volume, which were already reported in a previous study.^8^ Previous studies investigating neurological disease in adults have reported a positive correlation between choroid plexus and lateral ventricles volume,^12,28^ and our study adds to this knowledge by demonstrating a positive correlation between choroid plexus and lateral ventricles volumes in the pediatric population. However, the causal relationship between these volumes remains unclear. It is unknown whether a large volume of choroid plexus induces a large volume of lateral ventricles, or vice versa, or whether large ventricles lead to an overestimation of choroid plexus volume. In the case of CPH, it seems that overproduction of CSF induces a ventriculomegaly, resulting in hydrocephalus.

CPH is a rare pediatric disorder characterized by overproduction of CSF. VPS is the initial treatment for most cases, but almost all patients with CHP suffer from complications such as refractory ascites and may require further procedures such as BEC or removal of the choroid plexus.^6^ CSF production exceeding 2,000 mL daily has been reported,^6,29-31^ and it has been suggested that if CSF production does not exceed 1,000mL/day, hydrocephalus caused by villous hyperplasia of the choroid plexus can be resolved with VAS.^32^ In our study, one of three patients was treated only with VPS, while the other two patients producing over 2,000 mL CSF daily showed complications. Additionally, genes on 9p likely regulate growth of the choroid plexus and trisomy 9p and tetrasomy 9p have been associated with CPH.^24^ Two of three patients in our study showed trisomy 9p.

Currently, there are no clear radiological criteria for describing the dimensions of a normal choroidal plexus or CPH,^6,24^ resulting in subjective diagnoses. Our study provides normal values of the choroid plexus across different ages, which enable us to compared choroid plexus volumes among different ages using z-normalization. In the P2 case, after VPS treatment, the normalized intracranial volume and choroid plexus volume increased due to growth. However, the normalized lateral ventricles volume improved, indicating the effect of VPS. In the P3 case, BEC was added to VPS treatment. After the surgery, the normalized choroid plexus volume was reduced by approximately 20%, indicating the effect of BEC. Not only the normalized lateral ventricles volume but also the normalized intracranial volume decreased due to the unclosed anterior fontanelle. Our study objectively assesses the degree of CHP and the effect of surgery.

Our study has some limitations. First, we enrolled CT scans pediatric patients with head trauma, so there is a possibility that head trauma may have influenced our results. Post-traumatic hydrocephalus can develop weeks to months after brain injury, and its incidence varies widely (0.7-50%).^33^ However, we did not observe any cases of post-traumatic hydrocephalus in our collected CT scans Furthermore, as our results for intracranial volume and lateral ventricles volume are consistent with previous studies, we believe that the effect of head injury can be ignored. Second, the number of females in our study was smaller than that of males. To assess differences more accurately between genders, a larger sample size is needed. Third, we could not collect information about low-birth-weight infants, which may have affected the growth curves presented in our study. Fourth, while we assessed choroid plexus volume using CT images, previous studies used MRI to calculate volume,^11,13-15,27^ which may have led to overestimation or underestimation of choroid plexus volume, or incomplete segmentation. However, as one evaluator (HH) segmented the choroid plexus, we believe there is consistency. Fifth, we only investigated date from Japanese children, so there is a possibility that differences in human races may have affected our results.

However, as our results were similar to those obtained from populations other than Japanese,^8,15^ we believe these differences can be ignored. Finally, we only presented results for children up to 10 years of age, and growth curves for children over 10 years old remain unclear.

The growth curves we presented can aid physicians in objectively assessing hydrocephalus or choroid plexus-related disease. As pediatric hydrocephalus needs to be evaluated while accounting for head and intracranial ventricle growth, we believe that z-normalization is an effective way to assess pediatric hydrocephalus. Objective assessments have the potential to reveal factors related to complications after surgeries for CPH, which would be important for neurosurgeons planning to operate on patients with CPH.

## Conclusions

Our study using CT scans from Japanese pediatric patients with head trauma provides normal growth curves for intracranial volume, choroid plexus volume, and lateral ventricles volume. Choroid plexus volume and lateral ventricles volume demonstrate rapid increases until one year old, reaching approximately 1.5mL and 10mL, respectively, and plateau thereafter. Using z-normalization based on mean and SD values from these growth curves, we were able to objectively assess the degree of hydrocephalus in CPH and the effect of surgical intervention. Our findings will aid in the objective assessment of hydrocephalus and choroid plexus-related diseases, taking into account the growth of children, and promote scientific advancements in these fields.

## Abbreviations

BEC: bilateral endoscopic plexus coagulation
CI: confidence interval
CPH: choroid plexus hyperplasia
CSF: cerebrospinal fluid
CT: computed tomography
DICOM: Digital Imaging and Communications in Medicine
EVD: external ventricular drainage
F: female
M: male
m: month
mL: milliliter
MRI: magnetic resonance imaging
OSBP: one-stage bilateral plexectomy
SBP: staged bilateral plexectomy
SD: standard deviation
SWD: surgical wound dehiscence
VAS: ventriculoatrial shunt
VPS: ventriculoperitoneal shunt
y: year

## Credit author statement

**Hiroaki Hashimoto:** Conceptualization, Methodology, Software, Formal analysis, Investigation, Data Curation, Writing - Original Draft, Visualization, Project administration, Funding acquisition. **Osamu Takemoto:** Supervision **Keisuke Nishimoto:** Investigation. **Gento Moriguchi:** Investigation. **Motoki Nakamura:** Investigation. **Yasuyoshi Chiba:** Investigation, Resources, Supervision, Project administration.

## References

1. Dewan MC, Rattani A, Mekary R, et al. Global hydrocephalus epidemiology and incidence: systematic review and meta-analysis. J Neurosurg. 2018;130(4):1065–1079.

2. Raimondi AJ. A unifying theory for the definition and classification of hydrocephalus. Childs Nerv Syst. 1994;10(1):2–12.

3. Rekate HL. The definition and classification of hydrocephalus: a personal recommendation to stimulate debate. Cerebrospinal Fluid Res. 2008;5(1):2.

4. MacAulay N, Keep RF, Zeuthen T. Cerebrospinal fluid production by the choroid plexus: a century of barrier research revisited. Fluids Barriers CNS. 2022;19(1):26.

5. Mestre H, Mori Y, Nedergaard M. The brain’s glymphatic system: current controversies. Trends Neurosci. 2020;43(7):458–466.

6. Hallaert GG, Vanhauwaert DJ, Logghe K, et al. Endoscopic coagulation of choroid plexus hyperplasia: Case report. J Neurosurg Pediatr. 2012;9(2):169–177.

7. Tomita Y, Kameda M, Senoo T, et al. Growth Curves for Intracranial Volume and Two-dimensional Parameters for Japanese Children without Cranial Abnormality: Toward Treatment of Craniosynostosis. Neurol Med-Chir. 2022;62(2):89.

8. Cutler NS, Srinivasan S, Aaron BL, et al. Normal cerebral ventricular volume growth in childhood. J Neurosurg Pediatr. 2020;26(5):517–524.

9. Madhukar M, Choudhary AK, Boal DK, et al. Choroid plexus: normal size criteria on neuroimaging. Surg Radiol Anat. 2012;34(10):887–895.

10. İ mamoğlu H, Erkoç M, Şalk İ, et al. Evaluation of the normal choroid plexus size in adults with Magnetic Resonance Imaging. Cumhur Medical J. 2013;35(1):51–54.

11. Alisch JS, Kiely M, Triebswetter C, et al. Characterization of age-related differences in the human choroid plexus volume, microstructural integrity, and blood perfusion using multiparameter magnetic resonance imaging. Front Aging Neurosci. 2021;13:734992.

12. Lizano P, Lutz O, Ling G, et al. Association of choroid plexus enlargement with cognitive, inflammatory, and structural phenotypes across the psychosis spectrum. Am J Psychiatry. 2019;176(7):564–572.

13. Choi JD, Moon Y, Kim H-J, et al. Choroid Plexus Volume and Permeability at Brain MRI within the Alzheimer Disease Clinical Spectrum. Radiology. 2022;304(3):635–645.

14. Müller J, Sinnecker T, Wendebourg MJ, et al. Choroid plexus volume in multiple sclerosis vs neuromyelitis optica spectrum disorder: a retrospective, cross-sectional analysis. Neurol-Neuroimmunol. 2022;9(3).

15. Egorova N, Gottlieb E, Khlif MS, et al. Choroid plexus volume after stroke. Int J Stroke. 2019;14(9):923–930.

16. Hauptmann M, Byrnes G, Cardis E, et al. Brain cancer after radiation exposure from CT examinations of children and young adults: results from the EPI-CT cohort study. Lancet Oncol. 2023;24(1):45–53.

17. Muhm M, Danko T, Henzler T, et al. Pediatric trauma care with computed tomography—criteria for CT scanning. Emerg Radiol. 2015;22:613–621.

18. Astrand R, Rosenlund C, Undén J, for the Scandinavian Neurotrauma C. Scandinavian guidelines for initial management of minor and moderate head trauma in children. BMC Med. 2016;14(1):33.

19. Hashimoto H, Maruo T, Kimoto Y, et al. The association between diffusion-weighted imaging-Alberta Stroke Program Early Computed Tomography Score and the outcome following mechanical thrombectomy of anterior circulation occlusion. Interdiscip Neurosurg. 2023;33:101758.

20. Hashimoto H, Maruo T, Kimoto Y, et al. Burr hole locations are associated with recurrence in single burr hole drainage surgery for chronic subdural hematoma. World Neurosurg: X. 2023;19:100204.

21. Gao K, Kemp DE, Ganocy SJ, et al. Antipsychotic-induced extrapyramidal side effects in bipolar disorder and schizophrenia: a systematic review. J Clin Psychopharmacol. 2008;28(2):203–209.

22. Brix MK, Westman E, Simmons A, et al. The Evans’ Index revisited: new cut-off levels for use in radiological assessment of ventricular enlargement in the elderly. Eur J Radiol. 2017;95:28–32.

23. Toma AK, Holl E, Kitchen ND, Watkins LD. Evans’ index revisited: the need for an alternative in normal pressure hydrocephalus. Neurosurgery. 2011;68(4):939–944.

24. Furey C, Antwi P, Duran D, et al. 9p24 triplication in syndromic hydrocephalus with diffuse villous hyperplasia of the choroid plexus. Cold Spring Harb Mol Case Stud. 2018;4(5):a003145.

25. Farin A, Deutsch R, Biegon A, Marshall LF. Sex-related differences in patients with severe head injury: greater susceptibility to brain swelling in female patients 50 years of age and younger. J Neurosurg. 2003;98(1):32–36.

26. Arnaud K, Di Nardo AA. Choroid plexus trophic factors in the developing and adult brain. Front Biol. 2016;11(3):214–221.

27. Ricigliano VA, Morena E, Colombi A, et al. Choroid plexus enlargement in inflammatory multiple sclerosis: 3.0-T MRI and translocator protein PET evaluation. Radiology. 2021;301(1):166–177.

28. Tadayon E, Pascual-Leone A, Press D, et al. Choroid plexus volume is associated with levels of CSF proteins: relevance for Alzheimer’s and Parkinson’s disease. Neurobiol Aging. 2020;89:108–117.

29. Casey KF, Vries JK. Cerebral fluid overproduction in the absence of tumor or villous hypertrophy of the choroid plexus. Childs Nerv Syst. 1989;5:332–334.

30. Tamburrini G, Caldarelli M, Di Rocco F, et al. The role of endoscopic choroid plexus coagulation in the surgical management of bilateral choroid plexuses hyperplasia. Childs Nerv Syst. 2006;22(6):605–608.

31. Fujimura M, Onuma T, Kameyama M, et al. Hydrocephalus due to cerebrospinal fluid overproduction by bilateral choroid plexus papillomas. Childs Nerv Syst. 2004;20:485–488.

32. Britz GW, Kim DK, Loeser JD. Hydrocephalus secondary to diffuse villous hyperplasia of the choroid plexus: Case report and review of the literature. J Neurosurg. 1996;85(4):689–691.

33. De Bonis P, Anile C. Post-traumatic hydrocephalus: the Cinderella of Neurotrauma. Expert Rev Neurother. 2020;20(7):643–646.

