## Supplemental Table 1 for "Normal growth curve of choroid plexus in children: Implications for assessing hydrocephalus due to choroid plexus hyperplasia"

| Intracranial volume (mL) | 0m | 1m | 2-3m | 4-5m | 6-7m | 8-9m | 10-11m | 1y-1 | 1y-2 | 2y | 3y | 4y | 5-6y | 7-8y | 9-10y |
|---|---|---|---|---|---|---|---|---|---|---|---|---|---|---|---|
| Mean | 438.63 | 574.18 | 730.32 | 791.97 | 912.20 | 972.64 | 1023.18 | 1078.05 | 1139.16 | 1198.56 | 1241.70 | 1328.28 | 1300.65 | 1328.24 | 1393.35 |
| SD | 46.33 | 66.19 | 70.49 | 98.02 | 119.77 | 78.43 | 95.16 | 100.02 | 84.02 | 104.17 | 154.36 | 112.15 | 70.18 | 121.55 | 62.70 |

| Choroid plexus volume (mL) | 0m | 1m | 2-3m | 4-5m | 6-7m | 8-9m | 10-11m | 1y-1 | 1y-2 | 2y | 3y | 4y | 5-6y | 7-8y | 9-10y |
|---|---|---|---|---|---|---|---|---|---|---|---|---|---|---|---|
| Mean | 0.33 | 0.80 | 0.90 | 1.10 | 1.14 | 1.36 | 1.02 | 1.45 | 1.52 | 1.47 | 1.48 | 1.86 | 1.49 | 1.33 | 1.29 |
| SD | 0.25 | 0.49 | 0.45 | 0.35 | 0.41 | 0.52 | 0.22 | 0.49 | 0.58 | 0.51 | 0.41 | 0.40 | 0.60 | 0.58 | 0.48 |

| Lateral ventricles volume (mL) | 0m | 1m | 2-3m | 4-5m | 6-7m | 8-9m | 10-11m | 1y-1 | 1y-2 | 2y | 3y | 4y | 5-6y | 7-8y | 9-10y |
|---|---|---|---|---|---|---|---|---|---|---|---|---|---|---|---|
| Mean | 1.97 | 5.81 | 8.55 | 8.68 | 10.15 | 10.34 | 11.61 | 10.83 | 9.91 | 11.02 | 9.19 | 8.12 | 10.29 | 11.28 | 9.38 |
| SD | 0.88 | 4.29 | 3.48 | 4.22 | 6.05 | 4.68 | 4.71 | 6.03 | 5.55 | 5.57 | 6.21 | 2.22 | 5.83 | 9.36 | 3.98 |

Supplemental Table 1

The mean and standard deviation (SD) values calculated for each age-related cluster are presented. The phase labeled as “1y-1” corresponds to an age range of 12 to 17 months. The phase labeled as “1y-2” corresponds to an age range of 18 to 23 months. Figure 3A, 3B, and 3C was created using these mean values.

m, month; y, year.
